# Preferences and willingness-to-pay for expanded carrier screening programmes in the general population: An integrative systematic review and meta-analysis

**DOI:** 10.64898/2026.03.24.26349154

**Authors:** Myrabeth Yeo Juann, Yasmin Bylstra, Nicholas Graves, Jeannette Goh, Christina Choi, Shermaine Chan, Saumya Shekhar Jamuar, Robin Blythe

## Abstract

**Purpose:** To systematically review population preferences for expanded carrier screening programmes to inform service delivery and health policy.

**Methods:** PubMed, CINAHL, and Scopus were searched from 1995 to 2025 on carrier screening for autosomal or X-linked recessive genetic conditions across adult general populations. Included studies elicited preferences on attributes regarding the design or delivery of carrier screening programs. We extracted preferences for each attribute, mapped qualitative findings to these preferences, assessed risk of bias and performed meta-analysis on the willingness-to-pay for screening using Bayesian multilevel modelling. All findings are reported in 2024 USD.

**Results:** Thirty one studies, including 16 quantitative, 11 qualitative, and 4 mixed-methods studies were included. Participants expressed preferences for which conditions to include in ECS, joint vs individual screening, the value of information provided before screening, in-person over online counselling, type of healthcare provider, and preconception testing. Willingness-to-pay was right-skewed with 9% of participants not willing to pay any amount, a median of $107 and an interquartile range between $41 and $226. Most studies demonstrated a high risk of bias.

**Conclusions:** We report preferences of the general population regarding expanded carrier screening programmes, including suggested amounts for copayment if subsidised by the health system.

## INTRODUCTION

Carrier screening is a genomics-based screening tool aimed at identifying an individual’s and/or couple’s risk of transmitting a recessive genetic disease to the next generation. While carrier screening was historically performed in high-risk families, either based on ethnicity or family history, use of next-generation sequencing has allowed assessment of risk for multiple conditions in a single test, also known as expanded carrier screening (ECS)^1^. ECS can be applied to all individuals and/or couples seeking information on their reproductive risk, regardless of their perceived risk. It is estimated that 0.5-1.0% of couples may be at increased risk^2,3^, signifying the potential scale of impact of ECS on informed reproductive planning. While professional societies support ECS as a tool for reproductive planning^4–6^, it is also important to consider the perspectives of the general population for whom ECS is ultimately intended. To integrate ECS into routine preconception healthcare, the economics and implementation of service delivery should be considered^7^. This may include the willingness of couples to pay and the ability of health systems to provide these services cost-effectively.

Our research objectives were to understand the preferences of the general population for the implementation of ECS and to estimate willingness-to-pay for these services. We systematically appraised the literature to understand how future population-level ECS can be structured using an integrative review and conducted Bayesian meta-analysis to estimate willingness-to-pay for these programmes among the general population.

## MATERIALS AND METHODS

This systematic literature review was registered with the International Prospective Register of Systematic Reviews (PROSPERO) [CRD420250655372] in March 2025 and conducted in accordance with the Preferred Reporting Items for Systematic reviews and Meta-Analyses (PRISMA) reporting guidelines^8^.

All supplementary methods and results can be found in the appendices.

### Search strategy and study selection

A preliminary search was performed in PubMed to identify sentinel articles that studied general population preferences regarding the programme delivery of expanded carrier screening tests. We used the Systematic Review Accelerator tool to identify key terms and subject headings for the development of a comprehensive search string^9^. A systematic search was conducted across three databases (PubMed, CINAHL, and Scopus) for peer-reviewed journal articles published between 1995 and 2025 with no language restrictions. A detailed search strategy for each database is available in Appendix 1.

Peer-reviewed original research studies were included in this review if they met the following criteria: assessed carrier screening for autosomal or X-linked recessive genetic conditions; collected data from preconception, antenatal, and/or general populations of adults; and elicited preferences on attributes regarding the design or delivery of carrier screening tests. Studies were included if they elicited any preferences regarding single-gene tests or ECS attributes, even if this was not the study focus.

The following exclusion criteria were applied: only elicited preferences from specific interest groups, such as families of children with genetic conditions; restricted to views on morality of carrier screening; only evaluated tests for adult-onset conditions such as breast cancer; conducted tests for purposes other than family planning; and focused only on carrier screening for conditions presenting with mild morbidity.

The systematic review was conducted using Covidence^10^. Abstracts were deduplicated and screened first by title and abstract then by full text based on inclusion and exclusion criteria by two reviewers (MYJ and RB). Any disagreements between reviewers were resolved through discussion, where a third reviewer (SSJ) was consulted. Data were then systematically extracted into Microsoft Excel^11^. Studies conducted by the same authors that appeared to use similar surveys only had unique data extracted.

### Data extraction and analysis

We selected attributes of preferences derived from quantitative (survey) studies and incorporated qualitative data to contextualise our quantitative results. For quantitative studies, participant preferences for ECS delivery were extracted and grouped into attributes. Qualitative data were extracted if they corresponded to these attributes, deepening contextual understanding. Data extraction is shown in more detail in Appendix 2.

Quantitative studies reporting willingness-to-pay for ECS were analysed using Bayesian multilevel modelling. Studies typically elicited willingness-to-pay as a bounded value with uneven interval sizes and censoring (e.g., $0, $1 to $100, $100 to $1000, $1000 or over). Willingness-to-pay was therefore analysed as a latent variable, with the true value for each participant somewhere between the upper and lower bounds of the response. All willingness-to-pay responses were converted to 2024 USD using currency conversion rates as of 2 January 2024^12^. A participant-level dataset was constructed using the number of responses for each combination of lower and upper bounds. Each participant was assigned a random intercept for each study, with the exception of the same study ID for two papers by Van Steijvoort et al.^13,14^, which appeared to use the same willingness-to-pay question for two similar populations. Willingness-to-pay modelling is further described in Appendix 3. Model code, including model diagnostics, is available at https://github.com/robinblythe/ECS_WTP. All willingness-to-pay analysis was conducted in R version 4.5.1^15^, using the No-U-Turn-Sampler^16^ from the ‘brms’ package^17^.

A quality assessment of each study was conducted using the Risk of Bias in studies of Values and Utilities (ROBVALU) tool to determine the reliability of reported preferences^18^. The assessment was performed by one study author (MYJ) and validated by a second (RB). The ROBVALU tool focuses on the appraisal of methods for preference elicitation. The data extracted from each study in this review pertained only to preferences for ECS relevant to our study; therefore, our risk assessment was limited to bias of reported preferences, rather than the overall study quality. Risk of bias assessments are detailed in Appendix 4.

## RESULTS

The initial literature search identified 1,202 records. After removing 334 duplicates, 858 records underwent title and abstract screening. From these, 96 articles progressed to full-text review, with 31 records meeting inclusion criteria.

**Figure 1:**
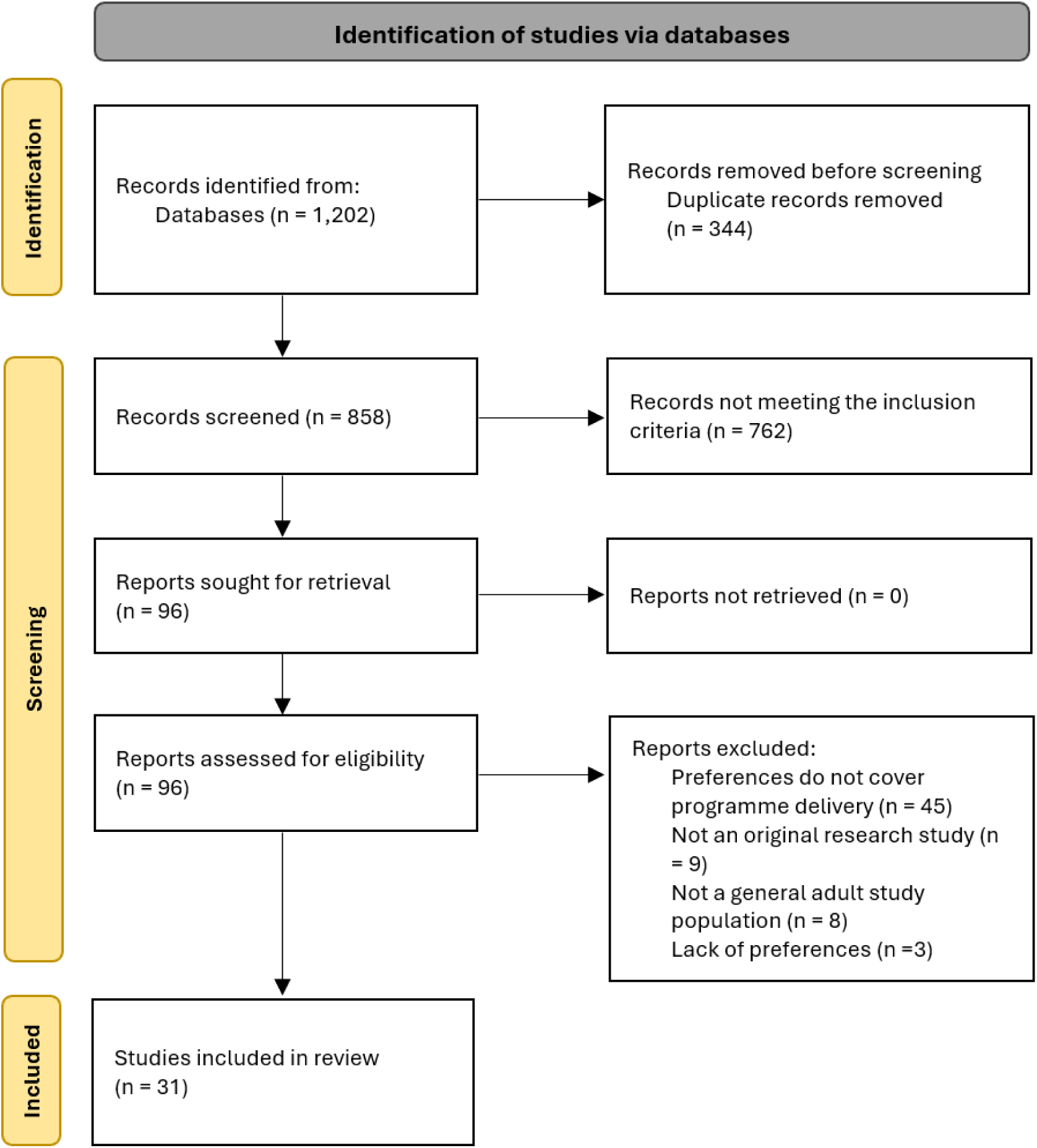
PRISMA flow diagram for study screening and selection.

Included records were dated from 1995 to 2025, with 16 quantitative^13,14,19–32^, 11 qualitative^33–43^, and 4 mixed-methods studies^44–47^. Studies included in this review were from the United States of America (USA), Australia, the Netherlands, China, Belgium, the United Kingdom, Slovenia, and Thailand. Of the included articles, 21 explored preferences for ECS, while 10 focused on carrier screening for a single or select group of recessive conditions. Included studies by study type are shown in Appendix 2.

We identified seven preference attributes in the reviewed studies: type of healthcare provider to offer ECS; details of ECS information provided to participants; screening method (joint vs individual), which genetic conditions to screen for; timing of screening (e.g., premarital or preconception); turnaround time for results; and willingness-to-pay. Studies used both inclusive (e.g., “select any that apply”) and exclusive (e.g., “select one”) survey questions.

### Risk of bias

Table 1 demonstrates the risk of bias across included studies. A detailed overall risk of bias assessment of the studies performed in this review is listed in Appendix 4. Generally, bias was attributable to the measurement instrument, mostly due to the incomplete representation of preference options or the use of selective data collection methods.

**Table 1:**
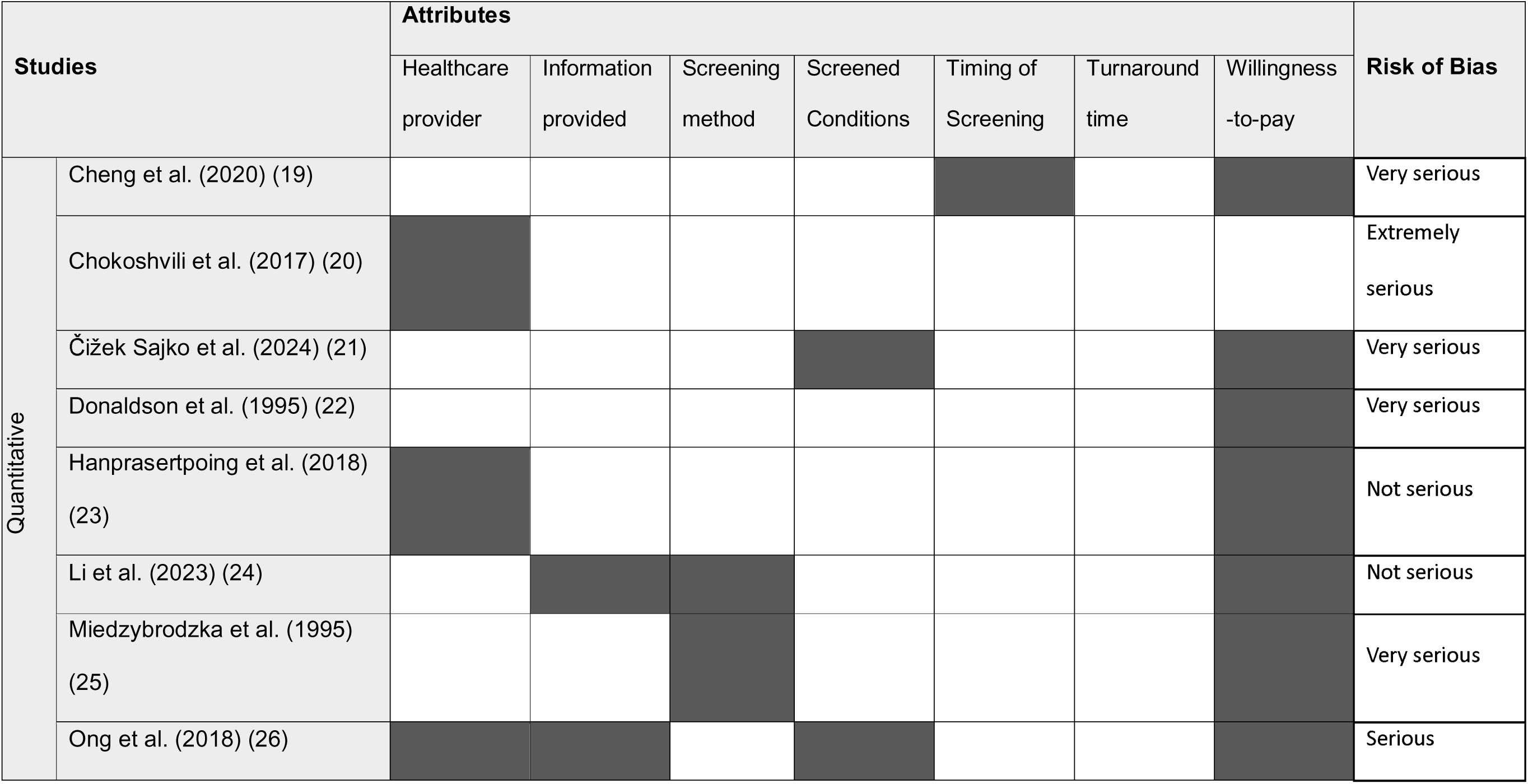
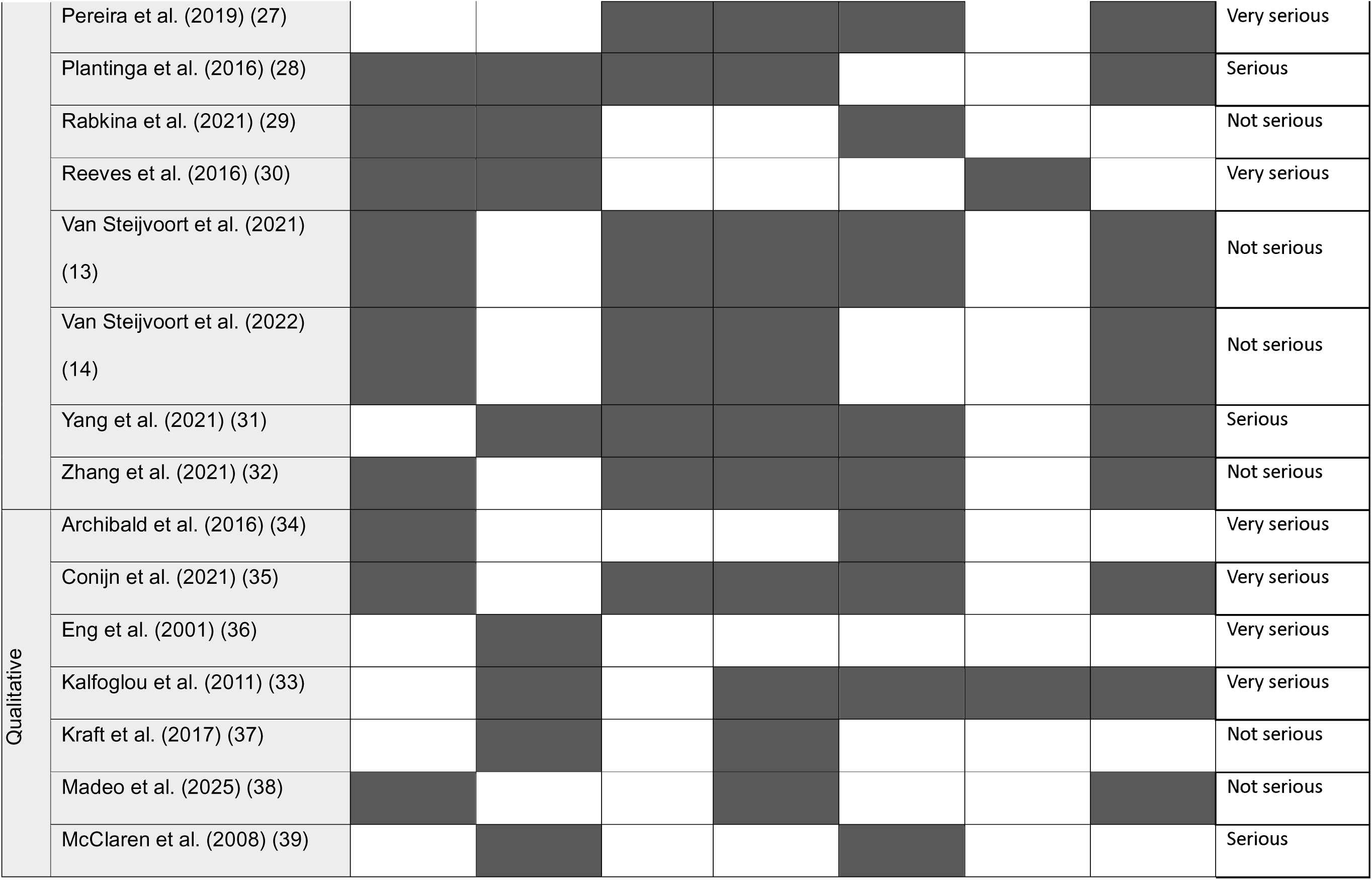
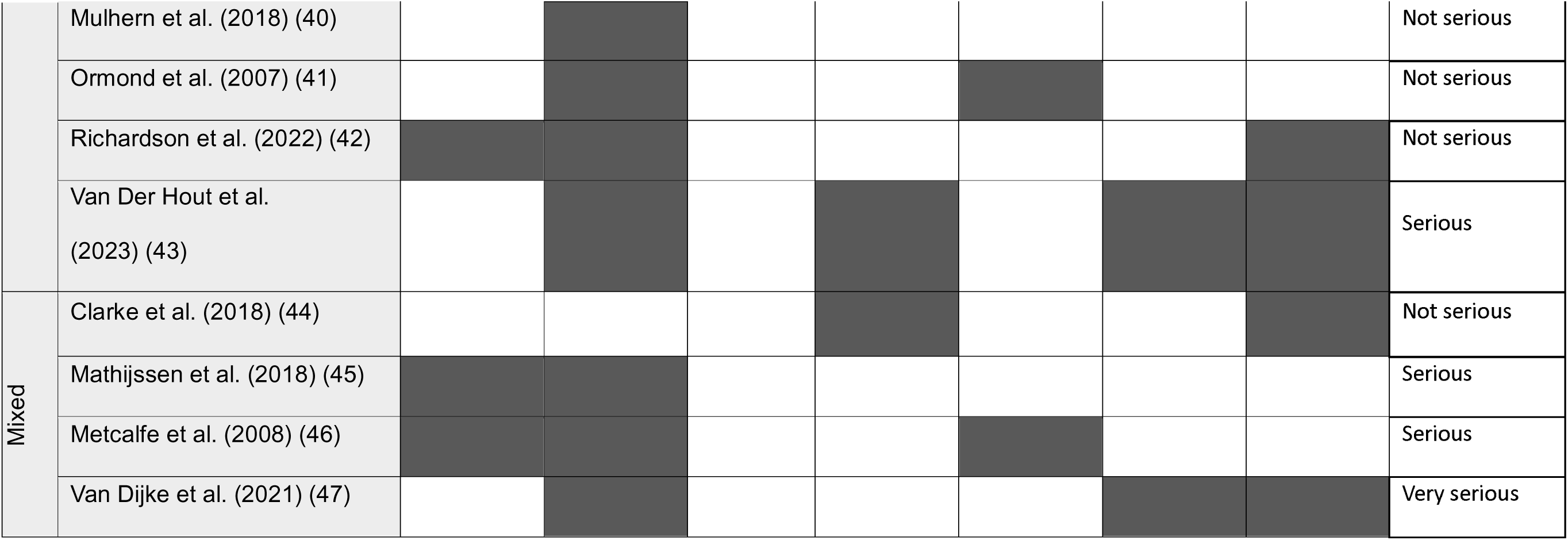
Summary of attributes reported and the overall risk of bias of studies included in this review. Shaded cells for each attribute indicate whether attributes were reported by a study, while unshaded cells indicate attributes not reported. The overall risk of bias for each study is shown in the rightmost column.

### Healthcare provider

Fifteen studies reported which healthcare provider participants preferred to offer and discuss screening; qualitative data from nine studies suggested that these varied by country and health system. Studies from the USA and EU reported strong preferences for obstetricians/gynaecologists (OB/GYNs) to handle both testing and education^29^. By contrast, 80% of Australian participants preferred their general practitioner to handle ECS, followed by their OB/GYN (42%)^26^. In China, 66% of participants preferred hospital-based screening delivery^32^, while Thai participants favoured provincial hospitals (74%) over private clinics^23^. Few studies assessed preferences for genetic counsellors, although Van Steijvoort et al. (2022) found that 65% of participants supported receiving care from counselling services^14^. Common reasons for selecting a type of clinical provider were accessibility^34^, the provider’s ability to pre-emptively recommend screening^35^, and the presence of an ongoing relationship with a clinician in whom participants had a high degree of trust^38^.

### Information provided

Results regarding information provided to participants were reported as preferences for in-person over online pre-test counselling and the amount of information participants wished to receive. Participants strongly supported pre-test counselling, with 94% of couples answering that pre-test consultations should always be provided^45^. Preferences were largely for in-person sessions over written or online materials, like websites or webinars, with 81%^29^ and 90%^36^ of participants preferring in-person or group counselling, respectively. In-person counselling (87%) was also strongly favoured over online counselling (7%)^47^. Richardson et al. (2022) found that these counselling sessions should allow sufficient time to address any concerns and recommend which tests were appropriate^42^. Participants generally preferred access to more, rather than less, information about ECS, including how serious or curable conditions were (88%) and what to do if the tests were positive (84%)^30^. Specific conditions tested (68%), technology used for screening (47%), and post-result pathways (44%) also helped participants decide whether to proceed with ECS^26^.

### Screening method

Preference between three screening methods were explored among nine studies: couple screening, in which both partners are screened together and receive a combined reproductive risk report; stepwise screening, in which one partner was screened and, if a carrier, the other partner was subsequently screened; and individual screening, in which both individuals in a couple receive a report of their individual carrier status. Three studies from China showed a strong preference for couple-based screening, with preferences ranging from 69% to 83% of participants^24,31,32^. By contrast, studies conducted in Europe and the USA reported preferences for individual or stepwise screening, with preferences ranging from 49 to 62%^13,14,25,27^. Miedzybrodzka et al. (1995) additionally reported that participants in the UK were willing to pay 63% more for stepwise over couple screening, and the minority who preferred couple screening were willing to pay 98% more for it^25^. Participants preferring individual screening believed it would be inconvenient to seek another couples test if they entered into a new relationship^35^. Women who were not yet in a relationship also wished to know their individual carrier status, as it would help make decisions about assisted reproduction. In a study of Orthodox Ashkenazi Jewish adults, participants wished to receive individual carrier results to reassure their children regarding the latter’s carrier status^33^.

### Screened conditions

Fourteen studies reported preferences for screened conditions, generally preferring the inclusion of ‘severe’ life-altering conditions and for a longer list of inclusions. Strong preferences were expressed for including conditions affecting the lifespan of children (92%) and those requiring full-time care (79%)^26^, conditions that are untreatable (85%) or cause foetal abnormalities (89%)^21^, conditions resulting in physical or intellectual disabilities (87%), and those leading to early death or shortened lifespan (77%)^32^. Expensive conditions to treat (53%)^32^ and non-health-related predispositions (19%) were considered less important^28^.

Overall, participants typically preferred more, not less, comprehensive screening. Qualitative studies reflected similar preferences for broader screening panels. However, participants expressed serious doubts about the inclusion of late-onset conditions in screening panels^35^. Providing categories of conditions to include in ECS, rather than a list of individual conditions, helped simplify the decision-making process regarding which conditions to include, including descriptions of living with different conditions^37^.

### Timing of screening

Twelve studies investigated preferences for carrier screening timing. Participants preferred ECS education to be provided prior to (51%) or when planning (19%) pregnancy^29^. Similarly, Yang et al. (2021) found that premarital (43%) and preconception (33%) screening were most preferred in comparison to childhood (14%), during school (5%) or prenatal (4.7%) screening. Participants generally felt that ECS should be routine for pregnant (92%) and fertility patients (88%)^27^, and should be offered to all couples who wish to have children^13^.

These preferences were shared across most qualitative studies^33,34,39,41^, which generally reported that preconception screening gave couples sufficient time to decide how to use their results. High school was mentioned as a possible setting to first introduce the concept of ECS^35^, and a handful of participants believed ECS could be normalised early as a universal component of healthcare^41^. Zhang et al. (2021) reported preferences for screening as late as the second trimester (41%), likely due to participants’ limited knowledge of ECS and misunderstanding of the test^32^. Cheng et al. (2020) reported that, when ECS was offered, more preconception women (71%) than pregnant women (61%) wished to participate^19^.

### Turnaround Time

Although four studies mentioned waiting times for ECS results, none explored what the preferred wait time would be or whether participants might be willing to pay more to expedite the process^30,33,43,47^. Wait times of 49 days^47^ and 100 days^43^ were both reported as too long. Studies reported that wait times directly led to increased stress^33^, especially if participants had previously suffered the loss of a child or pregnancy^43^. Participants also felt that turnaround time was considered less important than other attributes^30^.

### Willingness-to-pay

There were 4,671 willingness-to-pay responses for ECS among 8 quantitative studies^13,14,21,26,27,31,32,44^. Studies that reported willingness-to-pay for smaller screening panels were not included in this analysis^22–25,28,47^. Out of all responses, 405 (9%) were either not willing to pay for carrier screening, or the survey tool did not contain sufficiently granular estimates to determine whether participants would pay some small amount. The mean willingness-to-pay for ECS was $191 (SD $282) per person. The median was $107 [interquartile range (IQR): $41 to $226]. There was a 92% probability that willingness-to-pay for ECS was below $500.

2

Mean willingness-to-pay varied by study. The highest mean willingness-to-pay was observed in Europe (Slovenia and the Netherlands)^13,14,21^, while studies in China and Australia showed the lowest mean willingness-to-pay for ECS^26,31,32^. None of the above studies were explicit in the offered size of the panel, and mostly utilised the terms expanded carrier screening or reproductive genetic carrier screening when surveying willingness-to-pay. The bounded response variable (panel A), posterior density (panel B), and study-level means (panel C) are shown in Figure 2. Study-level summary statistics are included in Appendix 3.

**Figure 2:**
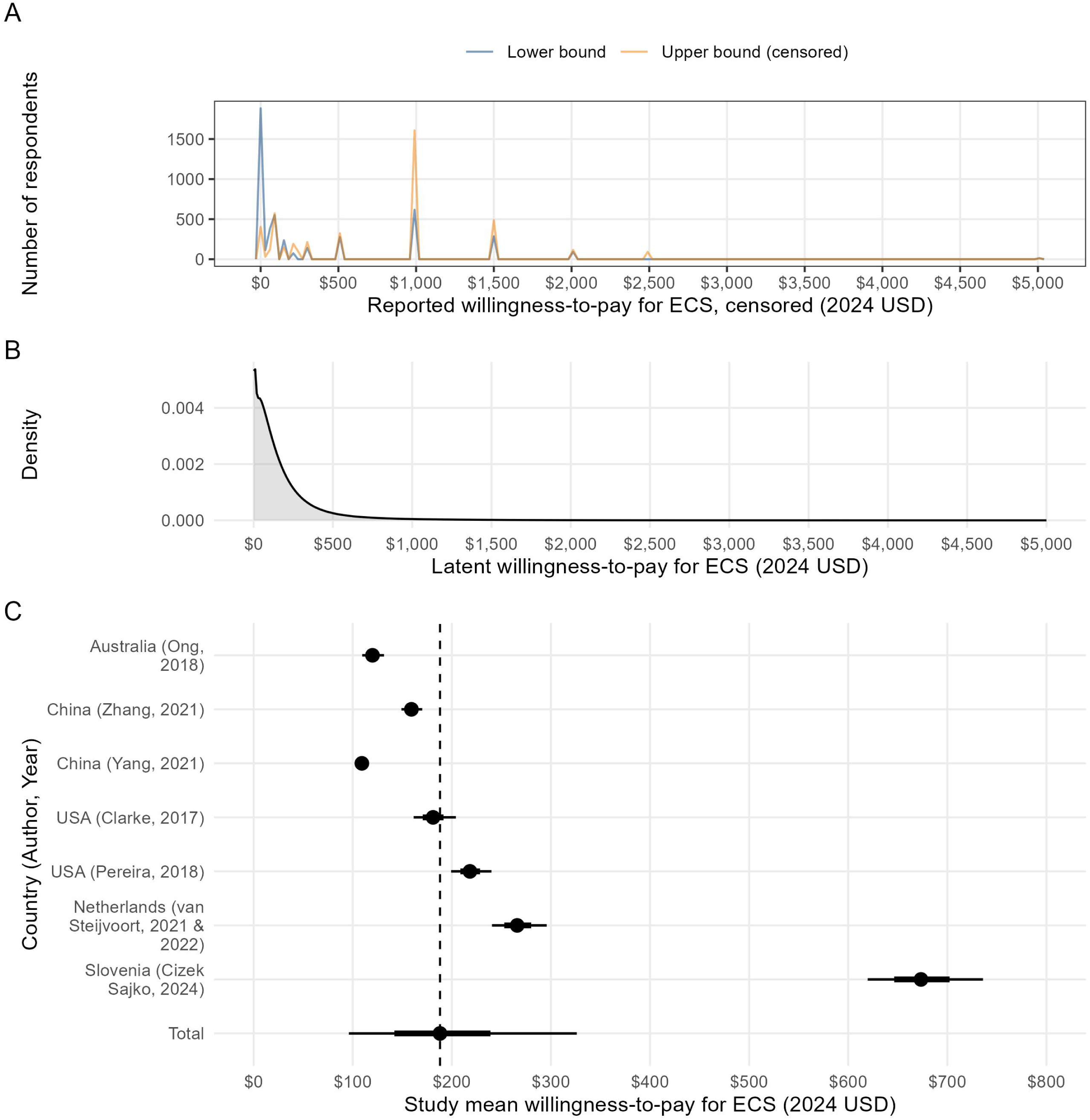
Willingness-to-pay for expanded carrier screening (ECS) in 2024 USD. Panel A displays the original survey responses, represented as lower and upper bounds in a smoothed density plot. Panel B represents the posterior distribution of the latent willingness-to-pay. Panel C shows study-level and overall mean willingness-to-pay for expanded carrier screening. The study mean is represented as a dot in the middle of each distribution, while the 66% and 95% distributions are represented by the shorter and longer lines, respectively, around each dot.

Several studies also evaluated the affordability of testing and whether participants believed screening should be covered by insurance^21–27,32,44^. In Thailand and Slovenia, 88%^23^ and 77%^21^ of participants, respectively, believed that screening should be government-funded. In the USA, Pereira et al. found that 54% of participants would only undergo ECS if covered by insurance^27^. Qualitative studies found that participants in Australia and the Netherlands believed that low income as a barrier to testing would lead to and perpetuate inequities^35,42^.

## DISCUSSION

In this systematic review, studies generally reported that the main benefit of carrier screening was to give participants enough time and information required to make an informed choice about family planning. Studies consistently reported that participants preferred screening to be available preconception or earlier and for pre-test counselling to be conducted in-person by a clinician or counsellor who remained available for any further queries. Participants generally requested sufficient information to understand the types of conditions included more broadly, but did not want to be overwhelmed with information about each condition. This was especially true of pregnant couples, who were already receiving information about their pregnancy. Trade-offs between more comprehensive ECS panels and panels that only included more severe conditions were not fully explored.

Based on these findings, we recommend that screening should be offered by trusted healthcare providers who can support individuals or couples throughout the screening process by providing or referring to counselling services. In line with study preferences, carrier screening should be available at least as early as preconception. Introducing screening to the premarital stage should be considered based on local cultural norms and the expertise of healthcare professionals. The impact of culturally and locally specific factors extends to the general population’s threshold of severity for conditions included in and whether screening is offered to individuals and/or couples.

Willingness-to-pay estimates were skewed by the use of unbounded upper intervals^21^. It is likely that many participants were unaware of the actual cost of ECS and may have calibrated their expectations according to survey’s framing and the assumed panel size. Differences across countries may be influenced by how prevalent ECS is within participants’ communities and by pre-existing knowledge of its cost within their healthcare system. Participants with a higher level of education and those with higher incomes were generally willing to pay more for ECS^13,14,26,32,44^. Eliciting willingness-to-pay is a complex process that can be conflated with ability-to-pay^48^; stated preferences can often differ depending on elicitation method^49^. We recommend that health systems apply preference elicitation methods like discrete choice experiments or examine demand curves for ECS to determine copayment levels acceptable to the local population. Health systems should also ensure that ECS affordability challenges do not widen disparities.

### Cultural and systemic factors

Preferences for screening methods were likely influenced by the relationship status of participants and the cultural norms of family planning in those settings. Single participants preferred individual results to inform alternative conception methods or a potential change in partners^14^; pregnant couples or those in committed relationships preferred couple-based screening^32^, as individual results were not relevant to their shared reproductive decision^35^. Couples in China^24,31,32^ also showed a preference for couple-based screening, while American and European participants preferred individual screening^13,14,25,27^. Ashkenazi Jewish participants who were more open to assisted reproductive technologies also preferred to receive individual test results or at least couple results with the genetic variant specified, differing from the standard disclosure policy of the Dor Yeshorim programme^33^.

Preferences for healthcare providers were likely informed by systemic factors. In Thailand and China, participants tended to prefer larger institutions like tertiary centres to provide ECS; by contrast, participants in Australia, the USA and Europe tended to prefer primary and secondary care modalities, including general practitioners and OBGYNs. This difference suggests that ECS uptake may be tied to broader perceptions of quality at different levels of local healthcare.

Tolerance for long turnaround times was rarely reported, but was regarded as undesirable given that major decisions rested upon the results. In the Dor Yeshorim program, carrier couples were discouraged from marrying, making longer waits especially stressful^33^ . Couples undergoing IVF or currently trying to conceive may also find long waits to be burdensome. While ECS programmes’ control over wait times is unclear, assessing willingness-to-pay for shorter waits could guide implementation.

### Research gaps and comparisons with existing literature

This review explored preferences of the general population for the attributes of ECS programmes intended to inform family planning. Van Steijvoort et al. (2022) assessed general population interest in ECS and found that participants’ intentions to undergo ECS were not always reflected in their subsequent decisions^50^; this suggests that the results of stated preference studies should be interpreted with caution. Ozdemir et al. (2022) reviewed studies of preferences for a wide range of genetic testing services^51^ and identified that test accuracy was an important factor for many participants; this was not generally explored in our included studies.

A significant research gap in our study results was the exploration of post-test decision-making and social support. The availability and utility of prenatal interventions may influence screening preferences. Following a positive test result, couples may wish to seek alternative reproductive options, which may vary between countries^52^. Singapore and Japan limit preimplantation genetic testing (PGT) to conditions classified as “severe”; conversely, the United States and Brazil have no regulations on PGT^53^. Dive et al. (2024) highlighted that the subsequent cost of any post-test interventions can also influence screening preferences^54^. We suggest that ECS programmes carefully consider how to reconcile differences in population preferences with legal and access-related barriers, which influence how programmes must ultimately be implemented.

An area of emerging research is how technology may support reproductive healthcare services. This has gained wider acceptance, particularly post COVID-19 pandemic, with patients finding digital tools like video consultations to be of equal quality to in-person consultations^55^. While most participants in our review preferred in-person counselling and information delivery, there may be validated digital solutions that can support ECS recipients. Given that healthcare providers have limited time, patients may feel they need more information than what can be covered within a routine consultation. Digital tools, like conversational agents or interactive educational platforms, could help bridge gaps in care and might improve autonomy, efficiency, and accessibility^56^. These, however, must be balanced against the need for culturally and contextually sensitive counselling services.

### Limitations

Studies included in our review used different methods to elicit preferences, including a wide variety of survey instruments. The use of exclusive or inclusive survey responses, for example, made it difficult to assess preferences on a similar scale. We suggest that discrete choice experiments or ranked-choice voting could mitigate some of these challenges, though we again note that stated and revealed preferences are likely to differ in this area^49^.

The decontextualised process of qualitative data extraction may have led to interpretations in our study that differed from those of the included study authors and from those of the study participants^57^. In the process of data extraction, we found that raw data were not available, making it impossible to reanalyse studies or identify whether we agreed with the interpretation of qualitative results provided by authors. In the case of survey responses, we suggest that future research make individual-level survey responses available, provided non-identifiability requirements are met.

Published studies in this review often recruited patients from healthcare provider practices or the internet. These populations may not be reflective of the broader population to whom ECS might eventually be offered. The perspectives of participants in our included studies, particularly regarding willingness-to-pay, may not reflect populations with language difficulties, financial limitations, or who are geographically remote. We therefore suggest adopting a more inclusive approach to localising ECS to the general population.

## Conclusions

In this study, we identified a range of preferences for ECS programmes that decision-makers should be aware of when implementing population-based screening. Our willingness-to-pay results could provide a basis for future economic evaluations seeking viable copayment amounts to offset health service costs, or as prior distributions in Bayesian models. It is important to note that although ECS participants are key stakeholders in determining program implementation, other stakeholders, including providers and funders, should also be considered. Future studies should consider how to balance participant preferences with systemic priorities necessary for practical implementation and evaluation.

## Supporting information

All supplementary methods and results can be found in the appendices.

## Data Availability

Data generated by this study are available at https://github.com/robinblythe/ecs_wtp.

## Data Availability

Data generated by this study are available at https://github.com/robinblythe/ecs_wtp.

## Acknowledgements

The authors would like to thank Adrian G Barnett, Frank E Harrell Jr and David C Norris for their suggestions for handling bounded and censored data using Bayesian methods. The authors would also like to thank Prof Ivy Ng, Prof Patrick Tan, Prof Jerry Chan, A/Prof Chan Yoke Hwee, and Prof Gareth Baynam for their guidance.

## Author Contributions

Conceptualisation: MYJ, NG, SSJ, RB. Data curation: MYJ, RB. Formal analysis: MYJ, RB. Funding acquisition: NG, SSJ. Investigation: MYJ, SSJ, RB. Methodology: RB. Project administration: SC. Software: RB. Supervision: NG, SSJ, RB. Validation: MYJ, YB, NG, SSJ, RB. Visualization: MYJ, RB. Writing – original draft: MYJ, YB, NG, JG, CC, SC, SSJ, RB. Writing – review & editing: MYJ, YB, NG, JG, CC, SC, SSJ, RB.

## Ethics Declaration

As this study only used information available in the public domain, institutional review board approval was not required.

