## Supplementary material for "Preferences and willingness-to-pay for expanded carrier screening programmes in the general population: An integrative systematic review and meta-analysis": All supplementary methods and results can be found in the appendices.

**Appendix 1 – Search strategy**

*Table A1: Search strategy for each of the three included databases: PubMed, CINAHL, and Scopus.*

| **Database** | **Search string** |
| --- | --- |
| PubMed | ((genetic[Title/Abstract]) OR (genomic[Title/Abstract])) AND  ((Preconception[Title/Abstract]) OR (carrier[Title/Abstract]) OR (ethnic*[Title/Abstract])) AND  (screen*[Title/Abstract]) AND  ((prefer*[Title/Abstract]) OR (perspective*[Title/Abstract]) OR (view*[Title/Abstract]) OR (attitude*[Title/Abstract]) OR (willingness to pay[Title/Abstract]) OR (choice*[Title/Abstract])) AND  (1995:2025[pdat]) AND  (journal article[Publication Type]) NOT  (systematic review[Publication Type]) |
| CINAHL | AB ( genetic OR genomic ) AND  AB ( preconception OR carrier OR ethnic* ) AND  AB screen* AND  AB ( prefer* OR perspective* OR view* OR attitude* OR willingness to pay OR choice* ) OR TI ( genetic OR genomic ) AND  TI ( preconception OR carrier OR ethnic*) AND  TI screen* AND  TI ( prefer* OR perspective* OR view* OR attitude* OR willingness to pay OR choice* )  Other ﬁlters applied: abstract available, peer reviewed, custom date range 1995:2025 |
| Scopus | ( TITLE-ABS-KEY ( genetic OR genomic ) AND  TITLE-ABS-KEY ( preconception OR carrier OR ethnic* ) AND  TITLE-ABS-KEY ( screen* ) AND  TITLE-ABS-KEY( prefer* OR perspective* OR view* OR attitude* OR willingness AND  to AND  pay OR choice* ) ) AND  PUBYEAR > 1994 AND  ( LIMIT-TO ( DOCTYPE , "ar" ) ) |

**Appendix 2 – Data extraction and study characteristics**

Quantitative studies (surveys) were summarised as the percentage of respondents selecting each answer to each question about ECS services. We sought to extract only survey responses that could inform attributes of an ECS programme administered to the general population. For example, data on the preferences of ECS healthcare provider were extracted; however, data on whether respondents thought carrier screening was moral were not extracted. Willingness-to-pay for ECS was extracted and analysed separately and described in the meta-analysis section.

Unlike quantitative studies, where we were able to directly extract the proportion of responses for each theme, qualitative studies reported the authors’ interpretations of the data, sometimes including quotations to emphasis key points. Study authors (RB and MY) read through the full text of each qualitative study and extracted author interpretations and representative quotations that corresponded with the pre-defined themes identified from the quantitative analysis. Rather than coding raw qualitative data from the included papers, which were often either not available or only available as quoted snippets, we deductively and descriptively mapped each finding or quote to a theme, reporting those that added insight to our extracted quantitative results. For example, when examining preferences for the choice of ECS service provider, we extracted data on the reasons why interview participants believed one provider might be superior to another.

We reported quantitative preferences as the preferences of respondents, while we reported qualitative preferences as the preferences of participants, to clarify any potential confusion about data sources for our results.

Study summaries are shown in Table A2 below.

*Table A2: Studies included in review by study type. Study aim is shown as reported by each study. Genetic testing is often reported by several names; generally, the name of the screening program employed by the study authors is listed.*

| **Study** | **Setting** | **Study Aim** | **Population** | **Sample size** | **Genetic test offered** |
| --- | --- | --- | --- | --- | --- |
| *Quantitative (survey) studies* | | | | | |
| Cheng et al. (2020) (23) | China | To assess knowledge and acceptance of ECS by pregnant and non-pregnant women in Hong Kong. | Women of reproductive age | 923 (623 pregnant) | ECS |
| Chokoshvili et al. (2017) (24) | Belgium | To explore attitudes of the Belgian general public on various aspects of medical genetic testing. | General population attending an annual cartoon festival | 1,182 | ECS |
| Čižek Sajko et al. (2024) (25) | Slovenia | To determine pregnant couples’ attitudes towards ECS, focusing on participation. | General adult population | 497 | ECS |
| Donaldson et al. (1995) (26) | Scotland | To establish the relative utility and willingness-to-pay for disclosure (stepwise) vs non-disclosure (couple) screening | Women attending antenatal clinic | 127 | Cystic fibrosis |
| Hanprasertpoing et al. (2018) (49) | Thailand | To examine patients’ knowledge, attitudes, and acceptance of antenatal thalassaemia screening and the factors influencing them. | Pregnant women | 1,006 | Thalassaemia |
| Li et al. (2023) (28) | China | To understand individuals’ attitudes and understanding of carrier screening. | Women of reproductive age | 1,673 | Spinal Muscular Atrophy |
| Miedzybrodzka et al. (1995) (29) | United Kingdom | To measure the value of stepwise vs couple-based screening. | Women randomised to stepwise or couple screening | 450 | Cystic fibrosis |
| Ong et al. (2018) (30) | Australia | To explore baseline levels of genetic knowledge and awareness of ECS, and to investigate factors influencing knowledge and attitudes to participation in future ECS programmes. | General adult population | 832 | ECS |
| Pereira et al. (2018) (31) | United States of America | To characterise current knowledge, attitudes, and desires for ECS. | General adult population | 521 | ECS |
| Plantinga et al. (2016) (32) | Netherlands | To investigate whether responsible implementation of population-based ECS is desirable and feasible. | Adult population with partners within reproductive age | 504 | ECS |
| Rabkina et al. (2021) (33) | United States of America | To better understand awareness, interest, and decision-making around ECS, and to assess women’s preferences for information and screening. | Women between 18 and 40 years, interested in having children, no family history of genetic condition | 260 | ECS |
| Reeves et al. (2016) (34) | USA | To assess preferences for level of detail provided to participants, to determine the importance of different pieces of information for decision-making, and to assess what sociodemographic factors influence these preferences. | University students | 693 | ECS |
| Van Steijvoort et al. (2021) (17) | Belgium | To explore perspectives of non-pregnant reproductive aged women regarding ECS and to identify respondent characteristics that influence these perspectives. | Women of reproductive age | 151 | ECS |
| Van Steijvoort et al. (2022) (18) | Belgium | To assess knowledge, attitudes and preferences for ECS. | General population between 18 and 49 years | 387 | ECS |
| Yang et al. (2021) (35) | China | To assess knowledge and attitudes of ECS among medical staff and general population. | Mix of general population and medical staff | 1,663 | ECS |
| Zhang et al. (2021) (36) | China | To explore attitudes, understanding, and possible misconceptions about ECS and identify factors affecting decision-making. | Adults aged 18-45 attending prenatal diagnosis visits | 888 | ECS |
| *Qualitative* | | | | | |
| Archibald et al. (2016) (38) | Australia | To explore stakeholder views about offering population-based genetic carrier screening for fragile X syndrome. | General adult population | 188 | Fragile X syndrome |
| Conijn et al. (2021) (39) | Netherlands | To study the perspectives of relatives of mucopolysaccharidosis type III (MPS III) patients and general population on ECS. | Relatives of MPS III patients and general adult population | 41 | ECS |
| Eng et al. (2001) (40) | USA | To identify the basis and characteristics of common genetic conditions in the Ashkenazi Jewish population and provide a framework for the development of future carrier screening programmes. | Ashkenazi Jewish adults | 42 (21 couples) | Genetic diseases common in Ashkenazi Jewish communities |
| Kalfoglou et al. (2011) (37) | USA | To better understand Orthodox Ashkenazi young adults’ knowledge, experiences, attitudes, and beliefs around genetic carrier testing. | Orthodox Ashkenazi Young Adults | 49 | ECS with or without Gauchers disease |
| Kraft et al. (2017) (41) | USA | To present evidence about patients’ decision-making processes in determining what ECS results to receive. | General adult population | 51 | ECS |
| Madeo et al. (2025) (42) | USA | To understand what informs women’s preferences for the timing of ECS and cancer predispositions, and determining the factors women consider when deciding upon a preferred clinician to oversee screening. | Women aged 20 to 35 attending OB/GYN clinics | 20 | Reproductive carrier and cancer predisposition screening |
| McClaren et al. (2008) (43) | Australia | To explore preferences for cystic fibrosis screening. | Adults at antenatal or pregnancy stage (mostly women) | 68 | Cystic fibrosis |
| Mulhern et al. (2018) (44) | USA | To understand patient perspectives on whether elevated risk of Parkinson’s disease should be divulged during Gaucher Disease screening. | Adults screened for β-glucocerebrosidase gene (non-carriers) | 75 | Gaucher disease |
| Ormond et al. (2007) (45) | USA | To assess how pregnant patients and their male partners perceive the consent process for screening, and whether generic consents would be acceptable alternatives. | General adult population | 41 | ECS |
| Richardson et al. (2022) (46) | Australia, Canada, NZ, UK, USA | To explore important outcomes to prospective parents accessing screening, and to explore the role of qualitative consultation with patients when developing a core outcome set. | Adults who have undergone ECS | 15 | ECS |
| Van Der Hout et al. (2023) (47) | Netherlands | To determine facilitators and barriers to decision-making with regard to ECS and reproductive options after a positive test result. | Adults in consanguineous relationships | 14 | ECS |

| *Mixed-methods* | | | | | |
| --- | --- | --- | --- | --- | --- |
| Clarke et al. (2018) (48) | USA | To measure participants’ willingness to pay for ECS. | General adult population | 227 | ECS |
| Mathijssen et al. (2018) (49) | Netherlands | To evaluate the views of a genetically isolated Dutch community regarding expanded carrier screening. | Dutch founder population | 319 | 4 conditions common to the Dutch founder population |
| Metcalfe et al. (2008) (50) | Australia | To develop a population screening model for Fragile X syndrome among non-pregnant women in primary care. | Non-pregnant women at family planning clinics | 320 (65 interviewed) | Fragile X syndrome |
| Van Dijke et al. (2021) (51) | Netherlands | To investigate experiences of past ECS participants. | Adults who have undergone ECS | 132 surveyed (16 interviewed) | ECS |

*ECS: Expanded Carrier Screening. RGCS: Reproductive Genetic Carrier Screening. OB/GYN: Obstetrician/Gynaecologist*

**Appendix 3 – Willingness-to-pay meta-analysis**

A Markov-chain Monte Carlo (MCMC) algorithm with 4 chains, 6,000 iterations per chain, and a burn-in of 3,000 iterations was specified, with thinning set to 3. A hurdle Gamma model was selected to allow for true $0 values, as some respondents may have moral objections to ECS and would refuse the service no matter the fee. We treated unbounded intervals (e.g., “$1000 or over”) as censored observations with no upper bound. We used a N(0, 2) prior for the intercept, a Γ(2, 1) prior for the shape parameter, a Beta(1, 1) prior for the hurdle parameter, and a half-Cauchy(0, 1) prior for the sigma parameter.

The hurdle parameter was 0.09 [95% credible interval (CI): 0.08 to 0.10], and the remaining posterior draws were Gamma distributed with µ = 5.29 [95% CI: 4.67 to 5.93] and α = 1.31 [95% CI: 1.25, 1.39].

*Table A3: Mean willingness-to-pay for expanded carrier screening by study, with 95% credible intervals around the mean willingness-to-pay value. All values reported as 2024 USD.*

| **Study** | **Country** | **Study mean WTP** | **95% CI lower** | **95% CI upper** |
| --- | --- | --- | --- | --- |
| Yang et al. (2021) | China | $109 | $104 | $114 |
| Ong et al. (2018) | Australia | $120 | $109 | $132 |
| Zhang et al. (2021) | China | $159 | $149 | $171 |
| Clarke et al. (2017) | United States of America | $181 | $161 | $203 |
| Pereira et al. (2018) | United States of America | $218 | $200 | $240 |
| van Steijvoort et al. (2021 & 2022) | Netherlands | $266 | $241 | $297 |
| Čižek Sajko et al. (2024) | Slovenia | $675 | $621 | $736 |

*WTP: Willingness-to-Pay. CI: Credible Interval.*

**Appendix 4 - Assessment of risk of bias and adjustments to the ROBVALU tool**

While this tool was designed for studies measuring values placed on health outcomes, it also proved useful for evaluating preferences for the characteristics of healthcare services, making it appropriate for our review.

The Risk of Bias in Values and Utilities (ROBVALU) tool contains four subdomains: selection of study participants, completeness of data, use of measurement instruments, and data analysis. The s*election of study participants* assessed whether the sampling strategy was appropriate for our target population of adults of reproductive age, addressing concerns of selection bias. The c*ompleteness of data* was evaluated to determine whether there was a risk of bias due to low response rates. *Use of measurement instruments* examined potential bias introduced during questionnaire or interview development and application, and the effect of potential bias on the reliability of preferences elicited. For example, bias may have been introduced through the framing of questions within a survey, priming participants to think and answer negatively about preferences for couple-based or stepwise screening. Lastly, in the context of our review, *data analysis* assessed whether studies reported their findings in a way that was comparable to other studies in the literature.

*Table A4: Risk of bias evaluated by the Risk of Bias in Values and Utilities (ROBVALU) tool.*

| Sub domains | Signalling Question | Adjusted severity |
| --- | --- | --- |
| Selection of participants | Was an appropriate study sample selected from the study’s sampling frame? | No |
| Completeness of data | Was the attrition rate sufficiently low to minimise the risk of bias? | Yes |
| Measurement Instrument | Was the instrument used to measure patient values and preferences in a valid and reliable manner? | No |
|  | Was the instrument used in the intended way? | No |
|  | Was valid representation of the outcome (health state/ preference) used? | Yes |
|  | Did the researchers check for understanding of the instrument? | Yes |
| Data analysis | Were the results analysed appropriately to avoid influence of bias and confounding? | Yes |

Each signalling question in the ROBVALU tool utilised a 4-point Likert-type scale with the options of “yes”, “probably yes”, “probably no”, and “no”, corresponding respectively to the risk of bias judgement of “not serious”, “serious”, “very serious”, and “extremely serious”. This risk of bias judgment was then performed for each sub domains followed by an overall study risk of bias as reported in Table 2. Questions 2 (attrition rate), 5 (validity of health state representation), 6 (face validity of instrument), and 7 (risk of bias/confounding due to analysis) of the ROBVALU tool were less relevant to our research objectives. Therefore, even if these signalling questions fetched “probably yes”, they were classified as “no serious risk of bias” instead of “serious risk of bias” to more accurately reflect its impact on the study’s quality.

Generally, we opted to classify the overall study risk of bias as “very” or “extremely serious risk of bias” if at least one preference attribute was clearly affected in their inclusion in our results. This means that these studies contained results that were considered less likely to reflect participants’ true preferences. For example, risk of bias from descriptive reporting of results was only high if study authors chose to collapse Likert scale responses. We interpreted the risk of bias with discretion and preferences considered unreliable by both reviewers were excluded from our analysis.

**Appendix 4 – Risk of Bias**

*Table A5: Study-specific risks of bias as assessed by the Risk of Bias in Values and Utilities (ROBVALU) tool.*

| Studies | | Selection of participants | Completeness of data | Measurement Instrument | Data analysis | Overall Risk of Bias |
| --- | --- | --- | --- | --- | --- | --- |
| Quantitative | Cheng et al. (2020) (23) | Not serious | Not serious | Very serious | Not serious | Very serious |
|  | Chokoshvili et al. (2017) (24) | Very serious | Not serious | Extremely serious | Extremely serious | Extremely serious |
|  | Čižek Sajko et al. (2024) (25) | Not serious | Not serious | Very serious | Not serious | Very serious |
|  | Donaldson et al. (1995) (26) | Not serious | Not serious | Very serious | Not serious | Very serious |
|  | Hanprasertpoing et al. (2018) (27) | Not serious | Not serious | Not serious | Not serious | Not serious |
|  | Li et al. (2023) (28) | Not serious | Not serious | Not serious | Not serious | Not serious |
|  | Miedzybrodzka et al. (1995) (29) | Not serious | Not serious | Very serious | Not serious | Very serious |
|  | Ong et al. (2018) (30) | Serious | Not serious | Not serious | Not serious | Serious |
|  | Pereira et al. (2019) (31) | Not serious | Not serious | Very serious | Not serious | Very serious |
|  | Plantinga et al. (2016) (32) | Serious | Not serious | Not serious | Not serious | Serious |
|  | Rabkina et al. (2021) (33) | Not serious | Not serious | Not serious | Not serious | Not serious |
|  | Reeves et al. (2016) (34) | Serious | Very serious | Very serious | Not serious | Very serious |
|  | Van Steijvoort et al. (2021) (17) | Not serious | Not serious | Not serious | Not serious | Not serious |
|  | Van Steijvoort et al. (2022) (18) | Not serious | Not serious | Not serious | Not serious | Not serious |
|  | Yang et al. (2021) (35) | Serious | Not serious | Not serious | Not serious | Serious |
|  | Zhang et al. (2021) (36) | Not serious | Not serious | Not serious | Not serious | Not serious |
| Qualitative | Archibald et al. (2016) (38) | Not serious | Not serious | Serious | Very serious | Very serious |
|  | Conijn et al. (2021) (39) | Not serious | Not serious | Very serious | Not serious | Very serious |
|  | Eng et al. (2001) (40) | Not serious | Not serious | Very serious | Not serious | Very serious |
|  | Kalfoglou et al. (2011) (37) | Serious | Not serious | Very serious | Not serious | Very serious |
|  | Kraft et al. (2017) (41) | Not serious | Not serious | Not serious | Not serious | Not serious |
|  | Madeo et al. (2025) (42) | Not serious | Not serious | Not serious | Not serious | Not serious |
|  | McClaren et al. (2008) (43) | Not serious | Not serious | Serious | Not serious | Serious |
|  | Mulhern et al. (2018) (44) | Serious | Very serious | Very serious | Not serious | Very serious |
|  | Ormond et al. (2007) (45) | Not serious | Not serious | Not serious | Not serious | Not serious |
|  | Richardson et al. (2022) (46) | Not serious | Not serious | Not serious | Not serious | Not serious |
|  | Van Der Hout et al. (2023) (47) | Serious | Not serious | Not serious | Not serious | Serious |
| Mixed | Clarke et al. (2018) (48) | Not serious | Not serious | Not serious | Not serious | Not serious |
|  | Mathijssen et al. (2018) (49) | Not serious | Not serious | Serious | Not serious | Serious |
|  | Metcalfe et al. (2008) (50) | Not serious | Not serious | Serious | Not serious | Serious |
|  | Van Dijke et al. (2021) (51) | Not serious | Not serious | Very serious | Not serious | Very serious |

Ten studies were rated as having no serious risk of bias, seven studies as serious risk of bias, twelve as very serious risk of bias, and one as extremely serious risk of bias.
